# LUNG ULTRASOUND FINDINGS IN MEXICAN PATIENTS WITH SARS COV2 INFECTION

**DOI:** 10.1101/2020.07.16.20146704

**Authors:** Luis Fernando Paredes Fernández, Iván Ilescas Martínez

**Author notes:** Corresponding Author: (LP). These authors contributed equally to this work.

## Abstract

In late December 2019, a new disease reported at the time by an unknown pathogen was reported, which was later found to be a new variant of coronavirus, now called SARS-CoV2. This new disease had a very rapid global spread, causing multiple deaths in a short time, and which led to putting the entire world on health alert. In patients who have this disease, they present bilateral opacities in frosted multilobar glass with peripheral distribution. Some authors have suggested the use of ultrasound at the point of care for its early recognition.

In this study, we evaluated the findings of lung ultrasound in 25 patients admitted to the General Hospital Dr. Enrique Cabrera, Mexico, with a diagnosis confirmed by RT-PCR of SARS CoV2. This small retrospective study suggests that artifacts like glass rockets with or without the Birolleau variant (White lung), confluent B-lines, thick irregular pleural lines, and variable size (subpleural) consolidations are typical findings of lung ultrasound in patients with COVID-19 pneumonia. The presence of these findings is useful when evaluating patients with suspected COVID-19. In resource-limited and austere settings where chest radiography, CT, and RT-PCR are not available or the response time is long, lung ultrasound performed by trained personnel can be an aid in the diagnosis of COVID-19.

## INTRODUCTION

On December 31, 2019, the WHO was alerted to the appearance of unknown cause pneumonia in Wuhan, China. The etiologic agent was identified on January 7, 2020, as the novel-Coronavirus 2019, which was later renamed as Coronavirus 2-Acute Respiratory Syndrome (SARS-CoV-2). Since its identification, the number of patients with the new coronavirus disease 2019 (COVID-19) continues to increase rapidly in Mexico and abroad1. On day 06/11/20 (GMT −07: 00h) have been reported globally 7,273,958 cases and 413,372 deaths 2, while in Mexico have been reported 129,184cases and 15,357 deaths 3.

The COVID-19 pandemic is seriously affecting healthcare in many regions of the world. This new species of coronavirus has a specific tropism for the lower respiratory tract but causes severe pneumonia in a low percentage of patients. However, the rapid spread of this severe infection during this pandemic is forcing large numbers of patients to be hospitalized. According to a Chinese cohort study, approximately 14% of COVID-19 patients developed a severe disease and 5% developed critical illness 4. Among hospitalized patients, approximately 20-42% of patients developed acute respiratory distress syndrome 5, According to the report data of the Mexican Ministry of Health, they are very similar in our country 3.

It is important to diagnose and treat pneumonia in patients with COVID-19. For the diagnosis, computed tomography (CT) of the chest is more sensitive than the reverse transcriptase-polymerase chain reaction (RT-PCR) in the diagnosis of COVID-19, and its findings also correlate with progression. and recovery from illness 6–8.

On CT, patients generally have bilateral multilobar ground-glass opacity (GGO) with a peripheral or posterobasal distribution, and CT lesions evolve with the greatest severity of visible radiological findings around day 10 of symptom onset 9. Despite its usefulness, CT is not available in many environments with limited resources. Additionally, the disinfection of the CT machine after use by a patient with suspected SARS-CoV-2 infection will result in delayed care for other patients requiring CT examination. It is in this operational context that lung ultrasound was proposed to evaluate patients with probable COVID-19 pneumonia10–12. COVID-19 pneumonia has peculiar characteristics and can be studied by lung ultrasound (LUS) in the early approach to suspicious patients13. There is a preliminary experience in the use of pulmonary ultrasound in the initial diagnosis of atypical pneumonia and this tool has been used in the diagnosis of influenza virus infection during its previous epidemic outbreaks, and now during the COVID-19 pandemic, its use has resurfaced14,15. Ultrasound signs are not specific when considered in isolation, but observing some aspects of vertical artifacts can improve the diagnostic power of ultrasound examination. Furthermore, the combination of ultrasound signs in the patterns described and their correlation with laboratory tests on different phenotypes of the disease can allow a reliable characterization and be helpful for triage and patient admission16.

## METHODS

### This study was a retrospective, observational study

Patients diagnosed with COVID-19 by the SARS-CoV-2 RT-PCR test for nasopharyngeal exudate were included and evaluated by lung ultrasound in different areas of Hospital General Enrique Cabrera from April to May 2020.

For several days, an investigator (L.P.) performed serial lung ultrasound on 21 patients diagnosed with SARS CoV2 infection. The stored ultrasound images were carefully reviewed by two researchers (L.P. and I.I.). The records of the electronic file were reviewed to determine demographics, comorbidities, paraclinical, and radiographic findings. Pulmonary ultrasound studies were performed with a Mindray M5 ultrasound equipped with a 2.5-6 MHz multi-frequency convex transducer, subsequently, an Esaote Mylab 6 ultrasound with a high resolution multifrequency convex transducer was used. Ultrasound studies were performed along the clavicular midline in the bilateral anterior chest wall and scapular line, and Interscapular regions in the posterior chest wall at the patient’s bedside by an experienced operator (LP) with the patients in the supine position (and in the lateral position if the patient’s conditions allowed it), 6 regions in each hemithorax were analyzed. The transducer was covered with a transparent probe cover, and the transducer and portable ultrasound device were cleaned with disinfecting wipes after each use, following a pre-established protocol.

We use standard lung ultrasound terminology, established by Professor Lichtenstein since 1989 at his training center (CEURF)17.

### RESULTS

A total of 21 patients were identified, of whom 81% were men, the average age was 48.7 years (SD ± 13.43), clinical and demographic features are described in Table 1 and Table 2.

**Table 1.** Clinical characteristics of the studied population.

|  | Yes | No |
| --- | --- | --- |
| Previous Tobacco Use | 9 | 12 |
| Comorbidities |  |  |
| Obesity | 11 | 10 |
| Type 2 Diabetes | 7 | 14 |
| Hypertension. | 6 | 15 |
| Chronic Kidney Disease | 2 | 19 |
| Others |  |  |
| *Obstructive Sleep Apnea.* | 1 |  |
| Symptomatology |  |  |
| Fever | 21 | 0 |
| Headache | 21 | 0 |
| Cough | 21 | 0 |
| Myalgia | 21 | 0 |
| Arthralgia | 21 | 0 |
| Dyspnoea | 20 | 1 |
| Odynophagia | 15 | 6 |
| Fatigue | 15 | 6 |
| Nasal Congestion | 9 | 12 |
| Chill | 9 | 12 |
| Sputum | 7 | 14 |
| Sickness | 5 | 16 |
| Hyposmia | 3 | 18 |
| Rash | 2 | 19 |
| Vomit | 2 | 19 |
| Anosmia | 1 | 20 |
| Dysgeusia | 1 | 20 |
| Conjunctivitis | 0 | 21 |
| Haemoptysis | 0 | 21 |

**Table 2.** Paraclinical Characteristics of the studied patients.

|  | General |  |  | Arterial Gasometry |  |  |  |  | Complete Blood Count |  |  |  |  | Liver Function Test |  |  |  |  | Coagulation Test |  |  |  |  |
| --- | --- | --- | --- | --- | --- | --- | --- | --- | --- | --- | --- | --- | --- | --- | --- | --- | --- | --- | --- | --- | --- | --- | --- |
| Case | Age (range) | Gender | SpO2 on admission | pH | PCO2 | PO2 | HCO3 | SO2 | WBC | NEUT | LYMPH | PLT | VPM | ALT | AST | LDH | PCT | CRP | D-Dimer | CX | PT | aPTT | INR |
| 1 | 60-70 | M | 75 | 7.5 | 27 | 84 | 19 | 97 | 5 | 3.6 | 0.8 | 273 | 8.7 | 68 | 57 | 567 | <0.5 | 7.5 | 356 | 95 | 11 | 25 | 1.1 |
| 2 | 30-40 | F | 94% | 7.4 | 25 | 64 | 16 | 92 | 7 | 6.1 | 0.6 | 130 | 9 | 21 | 35 | 207 | <0.5 | 10 | 3450 | 27 | 9.2 | 36 | 0.9 |
| 3 | 70-80 | M | 54% | 7.4 | 32 | 85 | 19 | 96 | 13 | 12 | 0.7 | 295 | 9.1 | 30 | 27 | 269 | 10 | 16 | 1310 | 278 | 15 | 34 | 1.5 |
| 4 | 40-50 | M | 69 | 7.5 | 29 | 48 | 20 | 85 | 13 | 12 | 0.6 | 422 | 6.6 | 33 | 49 | 722 | <0.5 | 31 | 3180 | 472 | 12 | 34 | 1.2 |
| 5 | 40-50 | F | 85 | 7.4 | 30 | 23 | 20 | 42 | 6.1 | 4.9 | 1 | 191 | 9.9 | 74 | 85 | 557 | <0.5 | 28 | 458 | 366 | 11 | 33 | 1.1 |
| 6 | 30-40 | M | 77 | 7.5 | 26 | 77 | 19 | 96 | 8.1 | 6.9 | 0.9 | 457 | 7.1 | 58 | 43 | 419 | <0.5 | 1.7 | 10831 | 45 | 12 | 37 | 1.2 |
| 7 | 30-40 | M | 82 | 7.4 | 31 | 54 | 64 | 87 | 7 | 5.4 | 1.1 | 313 | 7.2 | 38 | 56 | 847 | <0.5 | 25 | 1940 | 56 | 12 | 28 | 1.1 |
| 8 | 40-50 | M | 83 | 7.4 | 33 | 35 | 20 | 88 | 5.3 | 4.6 | 0.5 | 144 | 11 | 39 | 55 | 281 | <0.5 | 19 | 789 | 285 | 10 | 31 | 1 |
| 9 | 60-70 | M | 89 | 7.4 | 26 | 63 | 15 | 91 | 9.8 | 8.9 | 0.7 | 156 | 9.8 | 20 | 24 | 301 | <0.5 | 5.9 | 541 | 42 | 10 | 24 | 1 |
| 10 | 40-50 | M | 83 | 7.4 | 44 | 33 | 27 | 63 | 4 | 2.7 | 0.9 | 155 | 8.6 | 32 | 34 | 230 | <0.5 | 14 | 191 | 50 | 9.6 | 31 | 0.9 |
| 11 | 10- 20 | F | 88 | 7.4 | 34 | 46 | 23 | 89 | 8.1 | 7.2 | 0.7 | 204 | 9.4 | 21 | 24 | 366 | <0.5 | 21 | 643 | 85 | 9.8 | 25 | 1 |
| 12 | 60-70 | M | 92 | 7.4 | 33 | 61 | 21 | 91 | 11 | 9.4 | 1.1 | 270 | 8.5 | 46 | 54 | 513 | <0.5 | 23 | 661 | 44 | 12 | 40 | 1.2 |
| 13 | 60-70 | M | 65 | 7.1 | 90 | 61 | 25 | 78 | 21 | 17 | 2.4 | 300 | 8.7 | 13 | 28 | 477 | 0.5 | 7.5 | 4800 | 131 | 13 | 33 | 1.3 |
| 14 | 60-70 | M | 84 | 7.1 | 66 | 72 | 53 | 88 | 13 | 11 | 1.1 | 200 | 8.4 | 33 | 51 | 725 | 2 | 37 | 2643 | 266 | 16 | 27 | 1.6 |
| 15 | 60-70 | M | 90 | 7.4 | 27 | 67 | 18 | 93 | 14 | 14 | 0.2 | 458 | 8 | 25 | 21 | 533 | 10 | 15 | 1870 | 28 | 17 | 42 | 1.6 |
| 16 | 40-50 | M | 75 | 7.4 | 35 | 45 | 22 | 81 | 14 | 12 | 0.9 | 225 | 8.4 | 69 | 35 | 787 | <0.5 | 21 | 679 | 191 | 14 | 29 | 1.4 |
| 17 | 40-50 | M | 69 | 7.5 | 29 | 48 | 20 | 85 | 13 | 12 | 0.3 | 422 | 6.6 | 33 | 49 | 722 | 0.5 | 31 | 3180 | 472 | 13 | 34 | 1.2 |
| 18 | 40-50 | F | 75 | 7.4 | 31 | 35 | 21 | 70 | 8.1 | 7.2 | 0.6 | 354 | 8.6 | 69 | 65 | 492 | <0.5 | 19 | 430 | 213 | 12 | 37 | 1.2 |
| 19 | 40-50 | M | 83 | 7.5 | 28 | 68 | 20 | 94 | 6.5 | 4.9 | 1.1 | 171 | 8.6 | 24 | 27 | 249 | 0.5 | 10 | 100 | 79 | 12 | 26 | 1.2 |
| 20 | 50-60 | M | 72 | 7.3 | 53 | 54 | 29 | 85 | 19 | 17 | 0.8 | 421 | 7.4 | 115 | 47 | 560 | 2 | 4.2 | 720 | 130 | 12 | 31 | 1.1 |
| 21 | 30-40 | M | 31 | 7.2 | 49 | 50 | 20 | 76 | 17 | 16 | 0.6 | 99 | 8.2 | 43 | 34 | 950 | <0.5 | 28 | 4010 | 265 | 10 | 26 | 1 |
| Mean | 48.71 |  | 77 | 7.4 | 37 | 56 | 24 | 84 | 11 | 9.3 | 0.8 | 270 | 8.5 | 43 | 43 | 513 |  | 18 | 2037 | 172 | 12 | 31 | 1.2 |
| Standard error | 2.931 |  | 3.1 | 0 | 3.5 | 3.7 | 2.6 | 2.8 | 1 | 1 | 0.1 | 25 | 0.2 | 5.4 | 3.5 | 48 |  | 2.2 | 536 | 31 | 0.4 | 1.1 | 0 |
| Half | 46 |  | 82 | 7.4 | 31 | 54 | 20 | 88 | 9.8 | 8.9 | 0.8 | 270 | 8.6 | 33 | 43 | 513 |  | 19 | 789 | 130 | 12 | 31 | 1.2 |
| Mode | 42 |  | 75 | 7.4 | 26 | 35 | 19 | 85 | 8.1 | 4.9 | 0.6 | 422 | 8.6 | 33 | 24 | 722 |  | 31 | 3180 | 27 | 10 | 31 | 1 |
| Dev. Standard | 13.43 |  | 14 | 0.1 | 16 | 17 | 12 | 13 | 4.7 | 4.5 | 0.4 | 116 | 1.1 | 25 | 16 | 218 |  | 10 | 2456 | 142 | 2 | 5.1 | 0.2 |
| Minimum | 18 |  | 31 | 7.1 | 25 | 23 | 15 | 42 | 4 | 2.7 | 0.2 | 99 | 6.6 | 13 | 21 | 206 |  | 1.7 | 100 | 27 | 9.2 | 24 | 0.9 |
| Maximum | 72 |  | 94 | 7.5 | 90 | 85 | 64 | 97 | 21 | 17 | 2.4 | 458 | 11 | 115 | 85 | 950 |  | 37 | 10831 | 472 | 17 | 42 | 1.6 |

Likewise, all the included patients underwent radiological studies (20 Chest X-Ray and 1 thorax CT) in 66% of the patients we found the ground glass pattern, and in 85% of the patients, mixed density opacification in the peripheral lung. The rest of the radiological findings are collected in table 3. Abnormal findings were found on lung ultrasound in all patients (Table 4). All patients had characteristic glass rockets (five B-lines or more: Figure 1, Supplementary Videos 1 and 2), 14 patients presented the Birolleau variant (Figure 2, video 3 and 4), which has also been named as white lung, the waterfall sign, or light beam in the literature. This phenomenon is an extreme variant of ground glass where all of Merlin’s space (the space between the pleural line, the shadows of the ribs, and the lower edge of the screen) is hyperechoic 12. In 10 patients we found septal rockets (three or four B-lines between two ribs). All the studied patients presented confluent B-lines (Figure 3, Video 5), and thickened and irregular pleural lines were observed in 20 patients, while variable size (subpleural) consolidations (Figure 4) were observed in 19 of them (There are still supporters of the label subpleural consolidations. Detecting consolidations using ultrasound is possible only if they are subpleural. The term subpleural is therefore redundant, so we use the term in parentheses). Large consolidation was only found in one patient, and this patient had remained for more than a week with mechanical ventilation. No images of pleural effusion were found in any of the studied patients.

**Table 3.** Radiological characteristics of the studied population.

| Case | Ground Glass | Local irregular shading | Bilateral irregular shading | Interstitial abnormalities. |
| --- | --- | --- | --- | --- |
| 1 | Yes | No | Yes | No |
| 2 | Yes | No | Yes | Yes |
| 3 | No | No | Yes | Yes |
| 4 | Yes | No | Yes | No |
| 5 | Yes | No | Yes | Yes |
| 6 | Yes | No | Yes | Yes |
| 7 | No | No | No | No |
| 8 | Yes | No | Yes | No |
| 9 | Yes | No | Yes | Yes |
| 10 | Yes | No | Yes | Yes |
| 11 | No | No | Yes | Yes |
| 12 | Yes | No | Yes | Yes |
| 13 | Yes | No | Yes | Yes |
| 14 | Yes | No | Yes | Yes |
| 15 | No | No | Yes | Yes |
| 16 | Yes | No | Yes | Yes |
| 17 | No | No | Yes | Yes |
| 18 | No | Yes | No | Yes |
| 19 | Yes | No | Yes | Yes |
| 20 | No | No | No | No |
| 21 | Yes | No | Yes | No |
| Yes | 14 | 1 | 18 | 15 |
| No | 7 | 20 | 3 | 6 |

**Table 4.** Ultrasonographic characteristics in the studied patients.

| No. Progressivo | Septal Rodets | Glass Rodets | Broleaou Phenomen (Light Beam) | Confluent B lines | Irregular and thickened pleural lines | Small subpleural consolidations | Consolidations |
| --- | --- | --- | --- | --- | --- | --- | --- |
| 1 | No | Yes | No | Yes | Yes | Yes | Yes |
| 2 | Yes | Yes | Yes | Yes | Yes | Yes | No |
| 3 | No | Yes | Yes | Yes | Yes | Yes | No |
| 4 | No | Yes | No | Yes | Yes | Yes | No |
| 5 | Yes | Yes | Yes | Yes | Yes | Yes | No |
| 6 | Yes | Yes | Yes | Yes | Yes | Yes | No |
| 7 | Yes | Yes | No | Yes | Yes | Yes | No |
| 8 | No | Yes | Yes | Yes | Yes | Yes | No |
| 9 | Yes | Yes | Yes | Yes | Yes | Yes | No |
| 10 | No | Yes | Yes | Yes | No | Yes | No |
| 11 | No | Yes | No | Yes | Yes | No | No |
| 12 | No | Yes | No | Yes | Yes | Yes | No |
| 13 | No | Yes | No | Yes | Yes | No | No |
| 14 | No | Yes | Yes | Yes | Yes | Yes | No |
| 15 | Yes | Yes | Yes | Yes | Yes | Yes | No |
| 16 | Yes | Yes | Yes | Yes | Yes | Yes | No |
| 17 | No | Yes | Yes | Yes | Yes | Yes | No |
| 18 | Yes | Yes | No | Yes | Yes | Yes | Yes |
| 19 | Yes | Yes | Yes | Yes | Yes | Yes | No |
| 20 | No | Yes | Yes | Yes | Yes | Yes | No |
| 21 | Yes | Yes | Yes | Yes | Yes | Yes | No |
| Yes | 10 | 21 | 14 | 21 | 20 | 19 | 2 |
| No | 11 | 0 | 7 | 0 | 1 | 2 | 19 |

**Figure 1.**
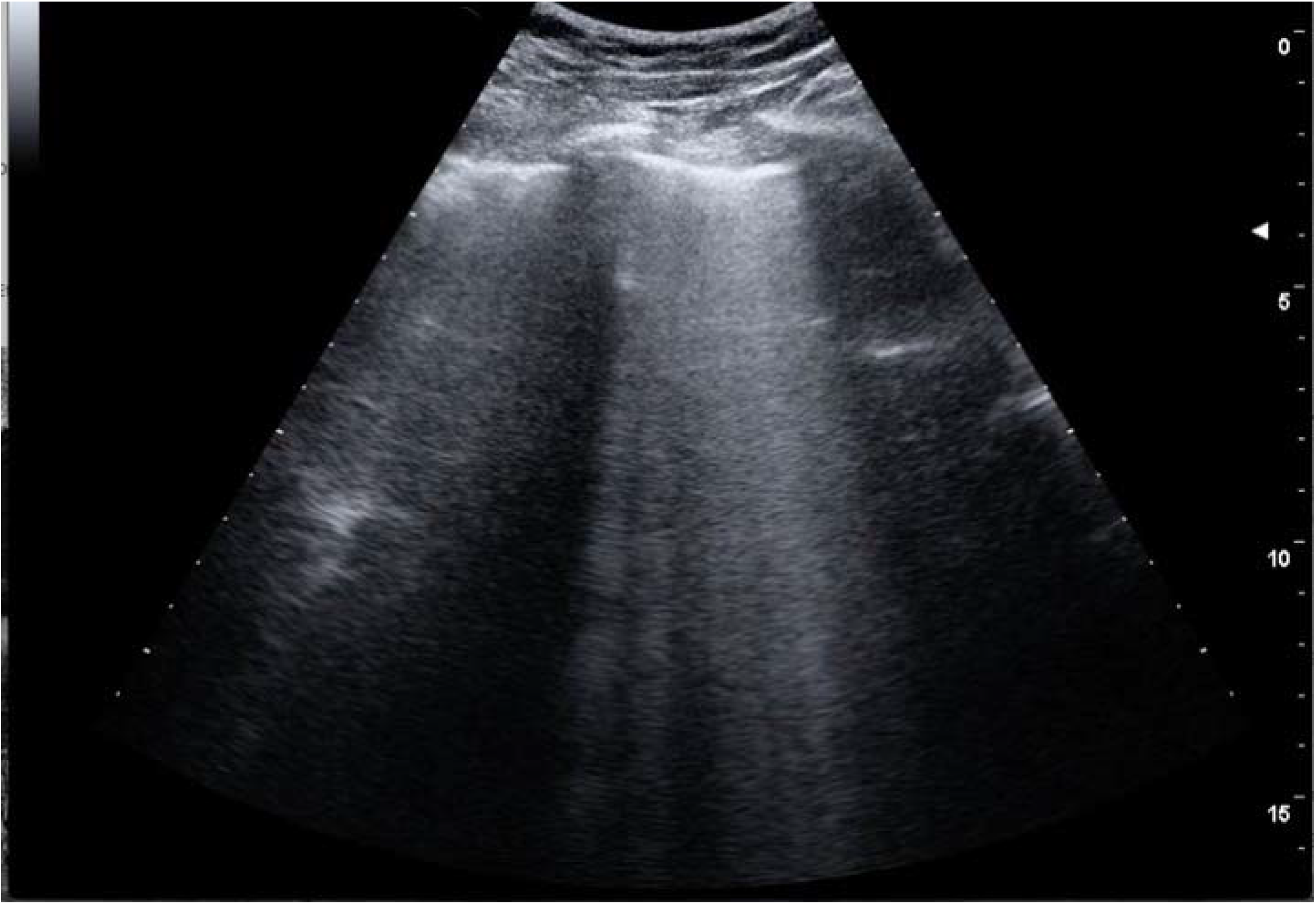
Glass Rockets

**Figure 2.**
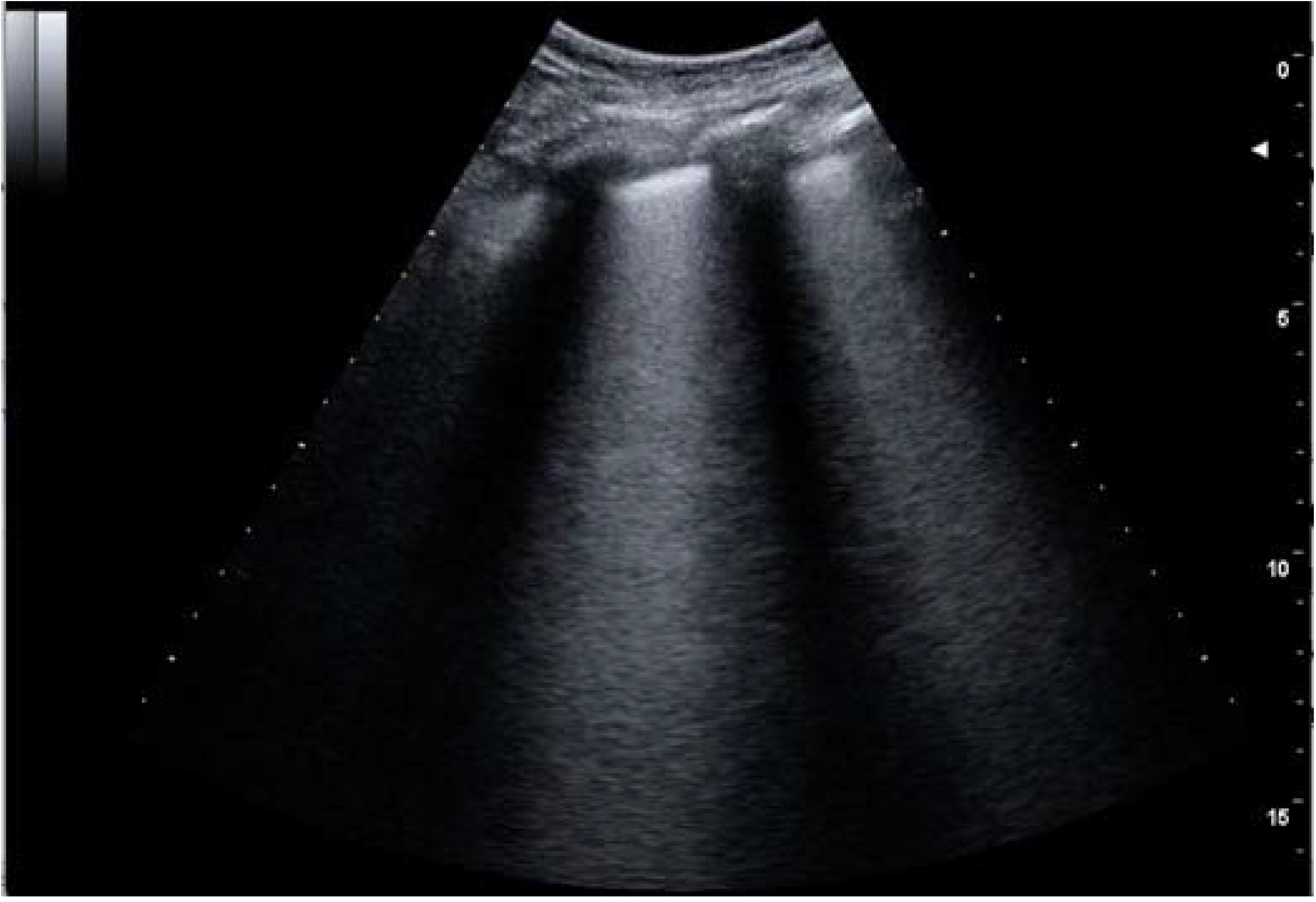
Birolleau phenomen (light beam).

**Figure 3.**
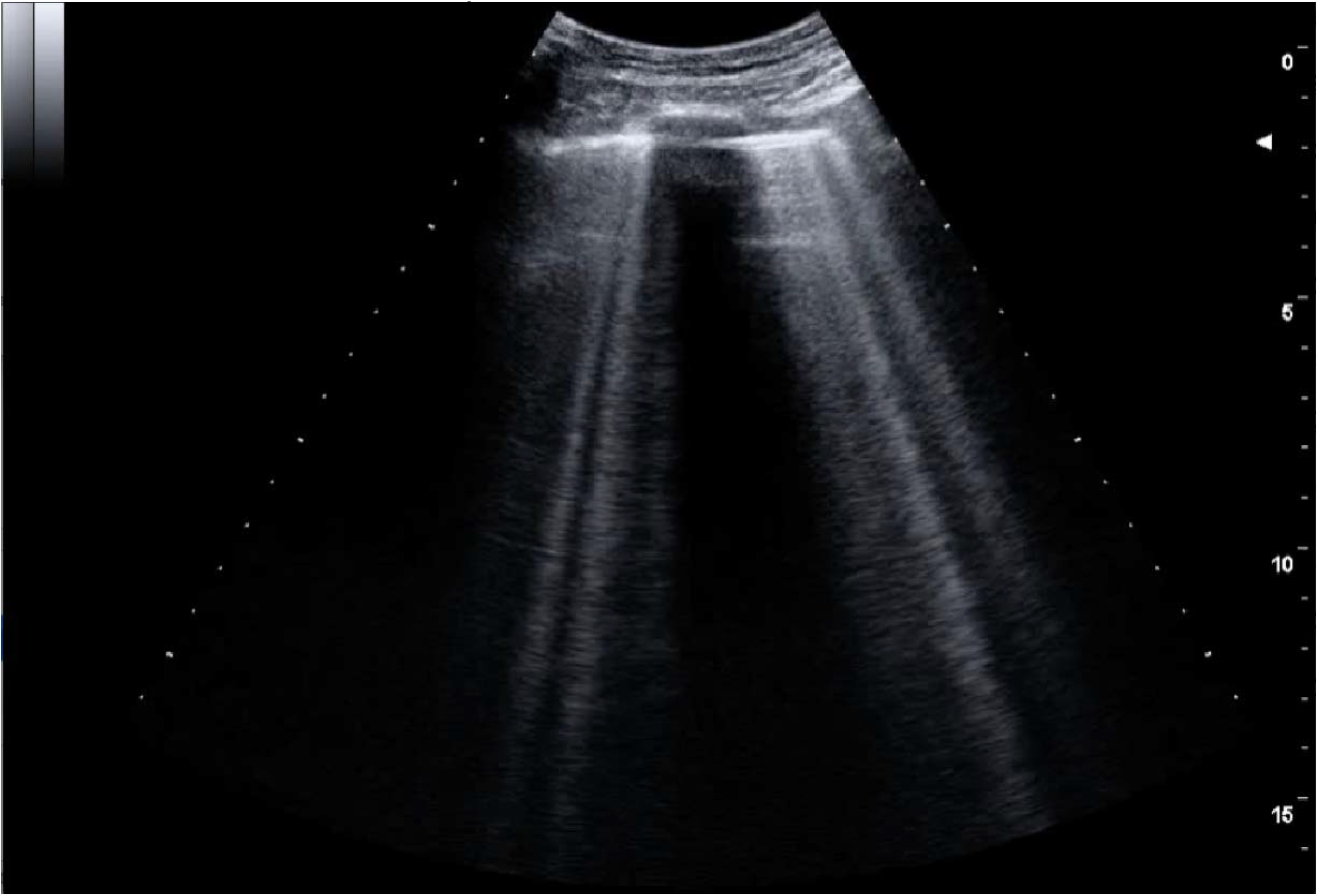
Confluent B lines.

**Figure 4.**
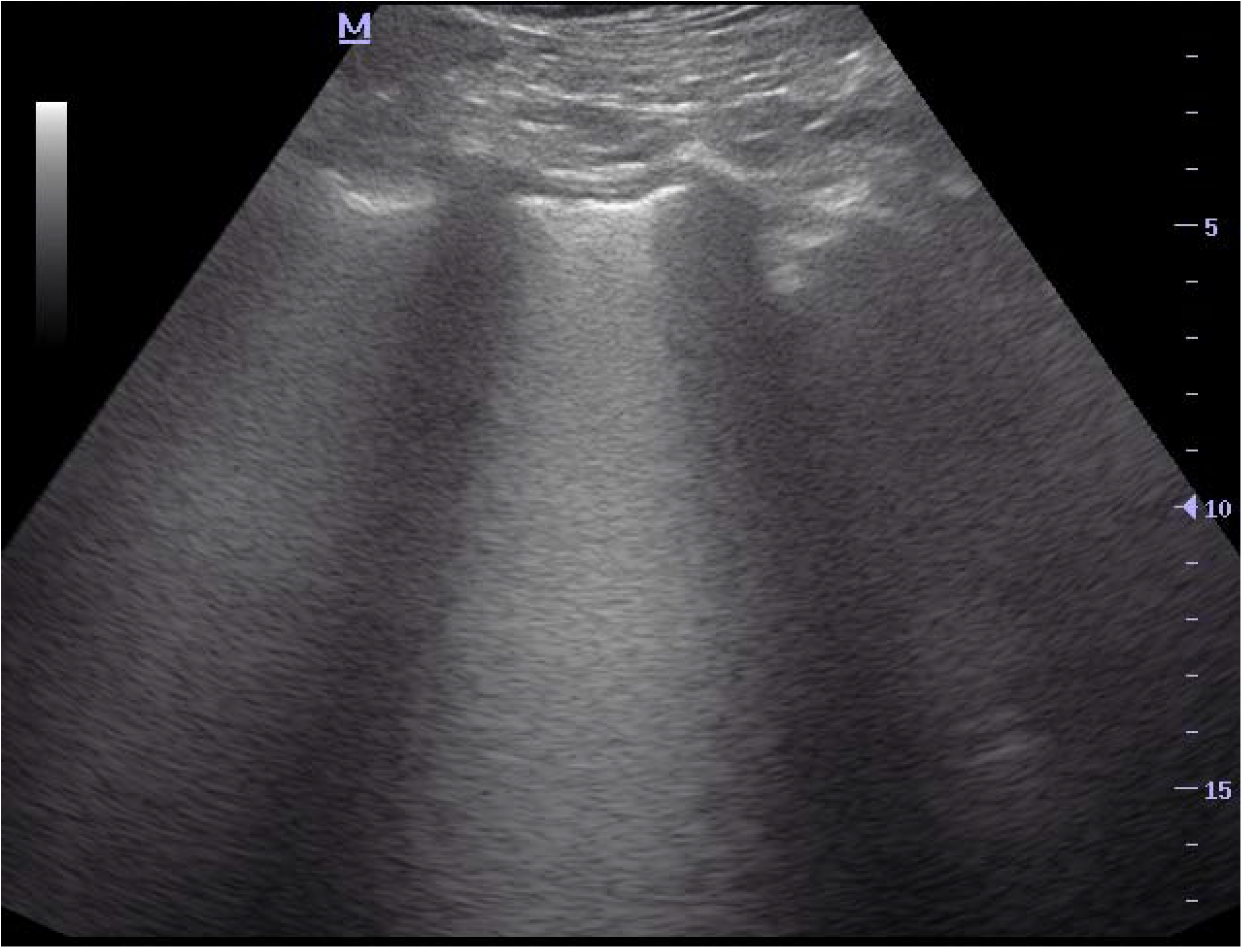
Thickened and irregular pleural lines.

In one patient, a chest CT scan was found in which mixed-density opacifications were reported in the pulmonary periphery, which on lung ultrasound were consistent with the appearance of the Birolleau variant.

## DISCUSSION

In our experience at the “Enrique Cabrera” General Hospital, we found that the vertical artifacts like septal rockets, glass rockets, or the Birolleau variant were visible in all patients with SARS-CoV-2 infection admitted to the different areas of the Hospital. These three ultrasound patterns are consistent with the areas of ground-glass opacity observed on CT, and in turn, are related to the presence of high-grade interstitial syndrome17, and although glass rockets and the Birolleau variant have also been observed in patients with pulmonary edema of cardiogenic origin, none of the patients studied was clinically diagnosed with decompensated congestive heart failure.

All the patients included in our study presented findings of pleural line thickening and irregularities, the addition of B lines and findings of pleural thickening suggest the appearance of a pleural inflammatory process and helps us to distinguish from those images that can occur in patients. with cardiogenic pulmonary edema.

Another important finding in the studied patients was the observation of variable size, usually small (subpleural) consolidations in 19 of them. In previous studies carried out in patients with Influenza pneumonia, the appearance of these images was reported, and the association between (subpleural) consolidations and confluent B-lines aided in distinguishing viral pneumonia from bacterial pneumonia 14,18.

Studies of ultrasound findings in patients with SARS-CoV-2 pneumonia are limited, but our findings are consistent with initial reports of the presence of “diffuse B-line pattern”, irregular and thickened pleural lines, confluent B-lines, and variable size (subpleural) consolidation. In our study, large consolidation was only observed in one patient, but the presence of large consolidation probably correlates with disease progression based on previous studies of CT findings in patients with COVID-19.

Since most patients with COVID-19 develop ground glass patterns of peripheral distribution that progress over time, lung ultrasound can probably detect most symptomatic patients with COVID-19 who require hospitalization. Glass rockets, confluent B-lines, thickened irregular pleural lines, and variable size (subpleural) consolidations are likely not specific to COVID-19 and can be seen in other conditions, such as other viral pneumonia and ARDS. However, these findings, particularly when combined and analyzed in context may be a decisive diagnostic aid during the COVID-19 pandemic when the pretest probability is high. Precisely, the high sensitivity of LUS for the detection of pulmonary involvement allows it to be a reliable monitoring tool for the assessment of patients with COVID-19.

Lung ultrasound has multiple advantages over chest X-Ray and chest CT in the diagnosis and treatment of patients with COVID-19. Lung ultrasound can be reasonably done at the patient’s bedside without exposing patients to radiation. Also, lung ultrasound is more sensitive than chest radiography in diagnosing alveolar-interstitial syndrome19. Lung ultrasound can detect lung lesions earlier than chest radiography when the lesions are adjacent to the pleura. Besides, the use of lung ultrasound in place of chest x-ray and chest computed tomography can reduce exposure of SARS-CoV-2 to healthcare workers, such as transportation personnel and radiological technicians, which can also help to mitigate the shortage of personal protective equipment experienced in many health facilities20.

Our study has some limitations. The main one is a retrospective study with a limited number of patients, as well as the relative scarcity of tomographic studies at the beginning of the epidemic in our institution. Future studies with a larger number of patients are needed to better assess the findings of lung ultrasound in COVID-19 patients and to evaluate the utility of lung ultrasound in treating patients with COVID-19. A more detailed evaluation with a previously validated scoring system, such as that used in intensive care for ARDS21,22, can provide prognostic information in patients admitted with COVID-19. Our examinations were carried out by a single expert sonographer (L.P.); therefore, these findings can be difficult for novice sonographers. It is also noteworthy that this study was conducted in patients who required admission to the internal medicine service. More studies are needed to characterize the sonographic findings of COVID-19 patients in other clinical settings.

## CONCLUSION

This small retrospective study suggests that artifacts like glass rockets with or without the Birolleau variant (White lung), confluent B-lines, thick irregular pleural lines, and variable size (subpleural) consolidations are typical findings of lung ultrasound in patients with COVID-19 pneumonia. The presence of these findings is useful when evaluating patients with suspected COVID-19. In resource-limited and austere settings where chest radiography, CT, and RT-PCR are not available or the response time is long, lung ultrasound performed by trained personnel can be an aid in the diagnosis of COVID-19.

## Data Availability

Does not apply.

## DECLARATIONS

### Ethics

The article was approved in its protocol form by the bioethics committee of the General Hospital Dr. Enrique Cabrera.

